# Public involvement to enhance care home research; collaboration on a minimum data set for care homes

**DOI:** 10.1101/2024.06.10.24308688

**Authors:** Anne Killett, Kerry Micklewright, Rachael Carroll, Gizdem Akdur, Emily Allinson, Liz Crellin, Kaat de Corte, Margaret Fox, Barbara Hanratty, Lisa Irvine, Liz Jones, Marlene Kelly Auburn Mere, Therese Lloyd, Julienne Meyer, Karen Spilsbury, Ann-Marie Towers, Freya Tracey, John Wilmott, Claire Goodman

## Abstract

**Introduction:** Information on care home residents is captured in lots of datasets (care home records, GP records, community nursing etc) but little of this information is currently analysed in a way that is useful for care providers, current or future residents and families or that realises the potential of data to enhance care provision. The DACHA study aimed to develop and test a minimum data set (MDS) which would bring together data that is useful to support and improve care and facilitate research. It is that utility that underscores the importance of meaningful public involvement with the range of groups of people affected. This paper analyses the public involvement of family members of care home residents and care home staff through a Public Involvement (PI) Panel.

**Objectives:** The objective for the PI activities was to consistently bring the knowledge and perspectives of family members and care home staff to influence the ongoing design and conduct of the DACHA study.

**Methods:** The bespoke methods of PI included a dedicated PI team and a PI Panel of public involvement contributors. Meetings were recorded and minutes agreed, resulting actions were tracked, and reflections on the PI recorded. A democratic, social relations approach was used to frame the analysis.

**Results:** A PI panel met 17 times. All meetings included both family members and care home staff. The public involvement deepened the research team’s understanding of the data environment in care homes, influenced the inclusion of quality of life and community health data in the pilot MDS and shaped research practices with care homes. Some panel members expressed personal and professional development from their involvement. Expectations of what the project could achieve had to be negotiated.

**Conclusions:** PI shaped the design and conduct of the DACHA study, grounding it in the needs and perspectives of people using and providing social care. Data research has a huge responsibility to accurately incorporate relevant public perspectives. There is an implicit assumption that records and data are objective and “speak for themselves” however there can be unintended consequences from introduction of new data requirements in practice.

**Patient or Public Contribution:** Public contributors to this manuscript include family members of older people living in care homes and staff of care homes. The wider study also involved as the public, older people living in care homes. Public contributors helped develop the project, contributed throughout the conduct of the study and some chose to be involved in preparing this manuscript.

## Introduction

The Developing research resources And minimum data set for Care Homes’ Adoption and use (DACHA) study aimed to develop and pilot a minimum data set (MDS) for care homes for older people in England to provide information to improve care and planning, and facilitate research, without overburdening care home staff. First a resident-level MDS was produced from data routinely collected by health providers, using a data linkage method which identified permanent care home residents aged 65 or older in NHS data sets. (1) This data was augmented with individually linked data from care homes’ digital care records. To develop the MDS, the study (2019-2024) drew on: data linkage opportunities between routine data sources beyond and within care homes; research evidence of other such minimum data sets internationally(2); measures used in care home research(3); implementation of care home research(4); a survey of care homes on data currently provided(5); national consultation activities(6-8); all underpinned by stakeholder involvement and engagement. This was a complex study awarded £2.2 million of funding, divided into 5 work packages (WPs) (see Figure 1), which took place over 4.5 years from November 2019. The 14 original collaborators came from 9 universities, the National Care Forum, The Health Foundation and the Alzheimer’s Society Research Network.

**Figure 1:**
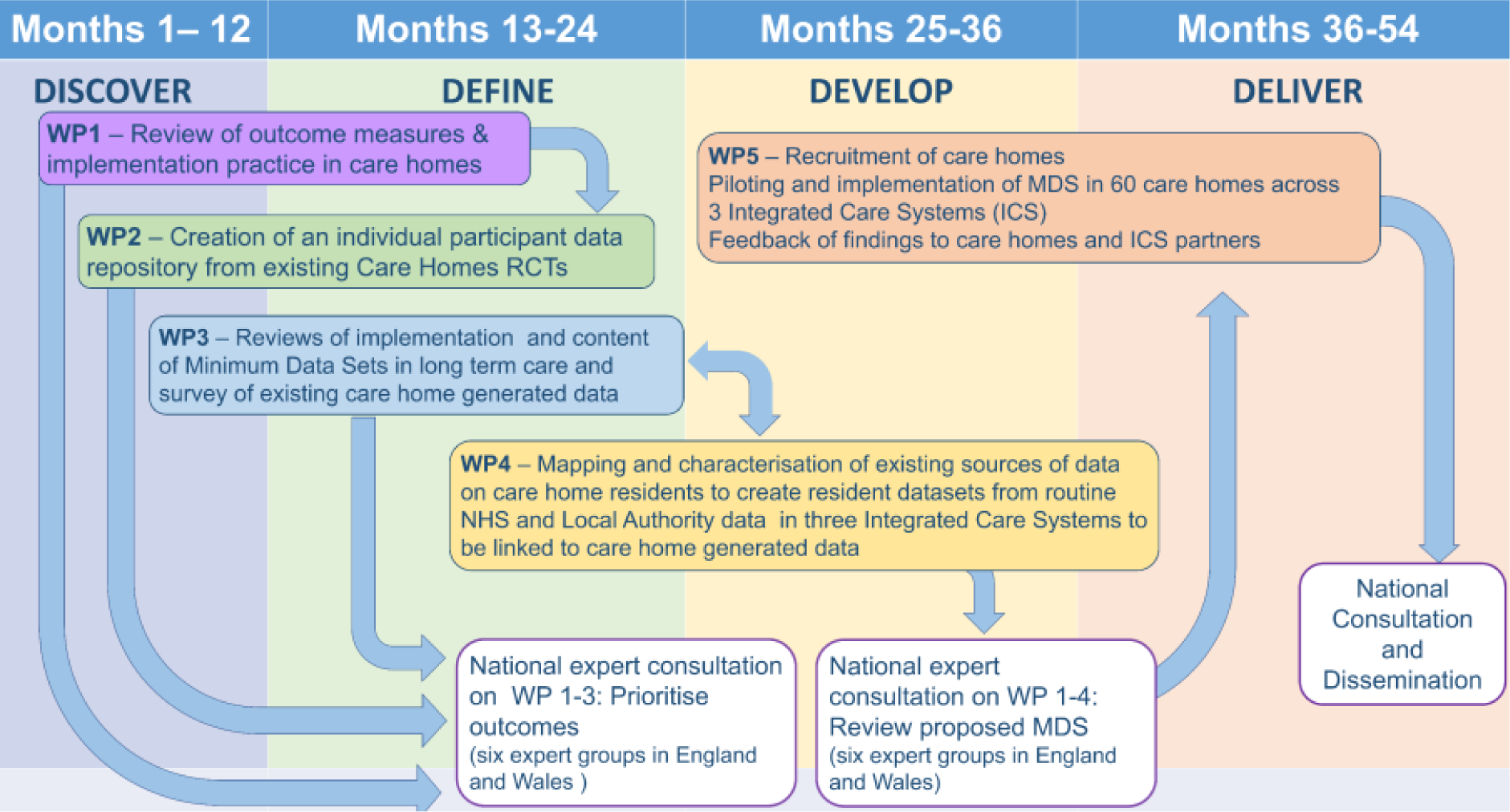
Diagram of the five Work Packages in the DACHA project.

**Figure 2:**
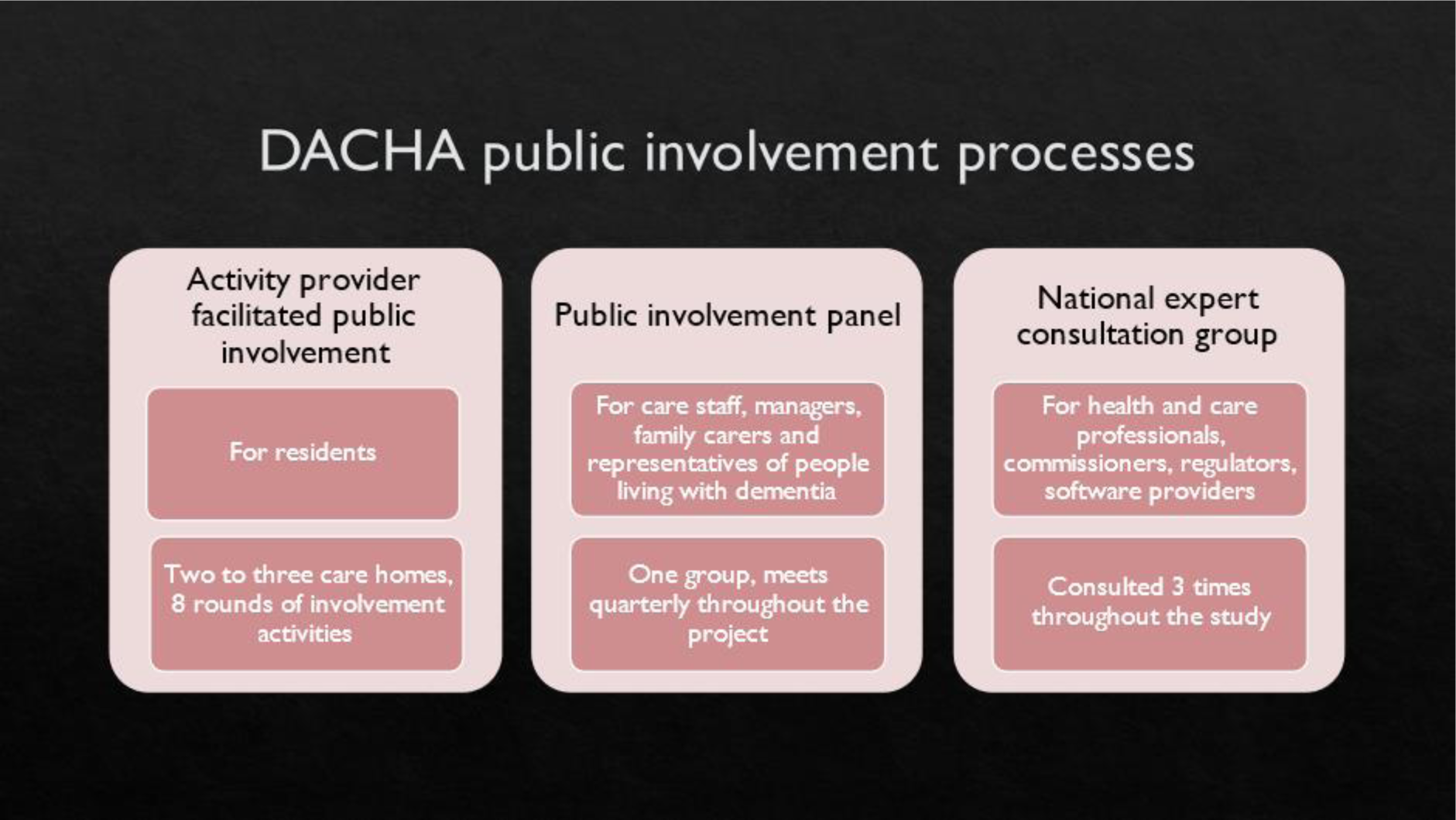
Three types of public involvement in the DACHA study.

There are important stakeholders in this enterprise, and the means of involvement or engagement need to be tailored to their interest, preference, area of existing knowledge and communication needs. Those most centrally affected by care home provision are the older people living in the care homes, and their families. Data recording activity or use of data to inform care has implications for those providing care: care staff and care home managers. Where there are practice implications there are related business and provision considerations for care home providers. Health care professionals and organisations are frequently involved in the care of older people in care homes (9). Local authority adult social services departments and the Care Quality Commission (CQC) are stakeholders who currently require data from care homes in their commissioning, quality assurance and regulatory roles. Care planning systems are moving from paper-based methods to digital systems, with a government target for 80% implementation in England by 2025 (10) therefore providers of digital care planning software are another relevant stakeholder group. Effective public involvement could reduce the risks of negative unintended consequences from findings and recommendations (11, 12).

Meaningful public involvement in care home research requires relationship building between researchers and public involvement stakeholders, and consideration of the differing perspectives and interests of stakeholders (13). Care home research programmes have involved patient and public involvement (PPI) representatives at each stage of the research cycle and in project management meetings (14) or in particular roles (15) but transparency about the extent, nature and influence of PI in published care home research is far from universal (16) and there have been calls for more discussion and debate of processes and evaluation (17). However, Edelman and Barron (18) argued for the evaluation of public involvement as a component of the research process rather than what they saw as a trend to evaluating public involvement in research as if it were a therapeutic intervention. Indeed such evaluation positions PI members as research participants, ‘othering’ them from more agentic roles in research (19). Rather than constructing and evaluating public involvement as an intervention it could be conceptualised as a social practice in which researchers and the public interact and power relations are considered (20). It is clear that research with care homes will be most effective with authentic co-production and active collaboration between researchers and care home representatives (4).

Frith (2023) argues that public involvement (PI) in applied health research is best understood as an attempt to make research more democratic, with potential to change both what is studied and the research processes, to broaden which knowledge is valued and who is involved in production of knowledge (21). PI and citizen science approaches have been compared, with some in the ‘science by the people’ tradition arguing for the place of local and contextual knowledge, indigenous knowledge, experiential knowledge and situated knowledge, alongside scientific knowledge, in consideration of technological risk and science policy (22). Care homes are a part of social care provision, which in the UK is argued to be in crisis and in need of a shift from a charity-welfare to a rights-based paradigm (23). Implementation of an MDS across Ontario inspired research with care workers who had few opportunities to contribute their knowledge at either organisational or policy level (24). Their experience of not having a voice was framed as epistemological violence. Fricker (25) proposed a concept of epistemic justice, and her description of hermeneutic injustice, where members of groups who do not have access to equal participation in generation of social meanings and are at a disadvantage when making sense of their social experience we argue can be applied to care home staff and relatives of care home residents. They are not equally included in the generation of social meaning about data and reporting requirements for care homes. Their experience is frequently marginalised. We therefore aimed to create a structure and social relations underpinned by democratic principles in which their knowledge could be recognised, valued and shape the knowledge developed by the project (25). The timing of involvement in relation to the stages of the project was important. With the findings of earlier work packages feeding into later work packages, we aimed for iterative engagement so that stakeholders could contribute to the developing understanding. Democratic principles underpinned the DACHA approach to public involvement, with the aim of enabling those least heard and most affected groups to contribute their own expertise and to advocate for others in the deliberation and decision making (21) of the project.

This paper presents an analysis of the involvement of care home staff and family members of care home residents in the DACHA project, using a social relations and democratic approach and reflection on examples of key effects over the four years of the study of the influence on the conduct and achievements of the study. Rather than separating out and evaluating or researching the PI component of the study we examine in context the challenges and demonstrate the effects of the involvement in building a project and outputs that are fit for purpose.

The DACHA project received ethical approval for distinct elements of the research project.

- Work Package 2: received ethical approval from Health, Science, Engineering & Technology ECDA – University of Hertfordshire (HSK/SF/UH/04185)
- WP3 national care home survey: received ethical approval from Health, Science, Engineering & Technology ECDA – University of Hertfordshire (HSK/SF/UH/04301)
- WP5 care home pilot: received ethical approval from the London Queen’s Square Research Ethics Committee (22/LO/0250).
- National consultation 2022: received ethical approval from Health, Science, Engineering & Technology ECDA – University of Hertfordshire (HSK/SF/UH/05009)
- National consultation 2023-24: received ethical approval from Health, Science, Engineering & Technology ECDA – University of Hertfordshire (HSK/SF/UH/05487)

Ethical review was not sought for this analysis of the PI process as it is not required for public involvement activity.

GRIPP2 reporting guidelines are followed(26).

## Materials and Methods

### Types of involvement

This section sets out the public involvement roles and structures in the project in order to set the context for the social relations between public involvement contributors and the research team. There were public involvement co-applicants, a Public Involvement team made of up public involvement members and researchers and PI member attending the core team meetings and representatives on the Study Steering Group. We had 3 tailored involvement and engagement processes to meet the needs of 3 different groups:

- Public Involvement Panel: Care home staff and family members of residents (discussed in this paper)
- Activity provider facilitated resident involvement: care home residents. (Reported elsewhere (27)).
- Consultation events: health and care professionals, commissioners, regulators, software providers along with broader representation from family carers, care staff and care home managers. (Reported elsewhere (6-8)).

### Public Involvement Panel

This paper examines the Public Involvement (PI) Panel. The PI Panel was a means of facilitating the involvement of family carers of care home residents, care home staff and care home managers. Five family carers, three care home staff and three care home managers formed the public involvement panel. On-line meetings were held quarterly throughout the project. Members of the PI team chaired, facilitated and took part in these meetings. Members of the wider DACHA team brought information from DACHA’s various work packages and asked questions of the panel so that the panel could influence the detailed design, implementation and interpretation of each of the work packages.

### Co-applicant roles

A family carer and a Director of The National Care Forum (NCF) were involved in planning the study from the early stages and were co-applicants in the application for funding of the study. Co-applicants met in hybrid meetings, combining in-person and on-line connection, and communicated through email, to design the project and respond to reviewer feedback throughout the funding application process. Once the project began co-applicants and researchers employed by the study were members of the Research Management Team (RMT) which met every other month online. Additionally, once COVID-19 restrictions were lifted the RMT met in person for a one or two-day meeting annually (2022-2024). In the second year of the project the family carer co-applicant resigned from the project and a person with experience as a family carer was recruited to join the PI team.

### Public Involvement team

There was a part-time senior research associate dedicated to the PI activity in the study (KM). Co-applicants responsible for public involvement in the project (AK, JM, LJ of NCF and family carer) met monthly online throughout the project to plan the ongoing involvement activities, and to act as a means of communication between the whole DACHA team and the public involvement panel and resident public involvement. The NCF involvement in the PI team and the PI panel brought broader care home representation and the policy context to inform discussions.

### Core research team

The core research team, which included the Chief Investigator, senior administrator and the senior research associates, met weekly for operational management of the project. Members of the PI team and a family carer with previous extensive experience of involvement in research and other public involvement roles joined this meeting once a month to coordinate the public involvement activities for the project, project responses to PI input and for the project to keep alert to family carer perspectives and concerns.

### Study steering committee

The study steering committee was chaired by a trustee of the former Residents and Relatives Association. Steering committee membership encompassed family carers, providers (business intelligence, software and care providers), health (commissioning and innovation), data governance, and data policy implementation. The committee aimed to meet a minimum of six times through the life of the study to act as a critical friend to the research team and provide advice, critical evaluation and guidance on all aspects of the study.

### Materials for analysing the process and impact of PI

Records of the PI activities included:

- Tracked actions of DACHA team members in response to input from the PI Panel (including feeding this back to Panel members) through RMT minutes and questionnaire sent to DACHA team.
- Minutes of PI Panel meetings
- Minutes of PI team meetings
- Notes of small group reflective discussions on PI (held at Research Management Team away day)
- Feedback from and discussion with members of the PI Panel (including use of UK Standards for Public Involvement, and leading to a reflective article by panel members(28))

These sources were read to triangulate and track developing impacts and identify themes (AK and KM) and emerging themes discussed and developed with MK, KM, RC, JM and then the rest of the co-authors.

### Recruitment

We recruited people to the PI panel through the Alzheimer’s Society Research Network, the National Care Forum, contacts with other care home researchers, existing university PPI groups and informal networks linked to the research team. We prepared a role description and information about remuneration. This was at the rate of £20 per hour for 2 hours of each meeting and 2 hours of preparation, offered to family carers and care home staff. We offered flexibility in how this could be paid; either a shopping voucher, payment as an individual (for which individuals would need to register with the university to fulfil UK employment law), or the care home could become a supplier to the university and invoice the university for the time of care home employees.

### Data recording – records of meeting and content

The PI panel meetings were held online using the Zoom™ platform. With the agreement of people attending the meeting, the meetings were recorded. PI team members wrote notes during the meeting which, with the recordings, were used to compile notes of the meeting, shared with panel members for their information and comments.

## Results

### Description of involvement activities

The PI panel met 17 times during the project (See Tables 1 and 2). Total attendances were: family carers 53; care staff 23; care home managers 24; DACHA PI team 70; DACHA WP teams 36 (see Table 1). Table 2 shows the meetings, attendees, agenda items, points emerging from discussions and how these were acted on in the study. Each of the five WP teams came to the panel at least twice, with three teams engaging four times with the panel. Panel members were sent an agenda and preparatory information two weeks before each meeting. To increase the accessibility of the information, PI team members fed back to research team members on draft information which was then edited before being sent to panel members. Panel meetings began in June 2020 when care home managers, staff, families and researchers were dealing with COVID-19. The COVID-19 outbreak had a massive impact on care homes, care home staff and on older people.

**Table 1:** PI panel meetings showing dates and numbers and roles of attendees.

| **Panel meeting number and date** | **Panel members – family carers** | **Panel members – care staff** | **Panel members – care home managers** | **PI team members** | **DACHA team members** |
| --- | --- | --- | --- | --- | --- |
| 1 30/06/2020 | 3 | 0 | 4 | 4 | 1 |
| 2 11/09/2020 | 3 | 2 | 3 | 5 | 1 |
| 3 05/02/2021 | 4 | 1 | 2 | 6 | 1 |
| 4 07/05/2021 | 3 | 3 | 3 | 6 | 3 |
| 5 06/08/2021 | 5 | 1 | 2 | 6 | 2 |
| 6 15/10/2021 | 4 | 2 | 0 | 5 | 4 |
| 7 05/11/2021 | 3 | 1 | 1 | 5 | 0 |
| 8 04/02/2022 | 3 | 1 | 1 | 4 | 4 |
| 9 06/05/2022 | 4 | 3 | 1 | 5 | 3 |
| 10 05/08/2022 | 2 | 2 | 0 | 4 | 2 |
| 11 04/11/2022 | 5 | 1 | 1 | 3 | 7 |
| 12 03/02/2023 | 3 | 1 | 1 | 4 | 1 |
| 13 05/05/2023 | 3 | 1 | 1 | 3 | 1 |
| 14 29/06/2023<br>meeting stopped through ill health |  |  |  |  |  |
| 15 04/08/2023 | 1 | 1 | 1 | 3 | 3 |
| 16 03/11/2023 | 3 | 2 | 1 | 4 | 1 |
| 17 01/03/2024 | 4 | 1 | 2 | 3 | 2 |

**Table 2:** Table of PI panel meeting agenda, key points and actions.

| Panel meeting number and date | Agenda items | Key points emerging | How key points taken account of in project |
| --- | --- | --- | --- |
| 1 30/06/2020 | Introduction to DACHA project | Will the two resident public involvement care home groups in Norfolk be representative? |  |
|  | Information about how the panel will work | When will the panel get information to look at so they can feedback on it? | Share research plan with key milestones for the project with the panel members. |
|  | Work package 2 – repository of data, is it reasonable to re-use original participants' data? | Panel members in favour of reusing data. Advised raising with ethics committee for advice. | Advice sought from ethics committee that originally approved a trial included in the repository. The view of PI panel that panel members were in favour of reusing data was shared with ethics committee. This trial is now included in the Trial repository. |
|  | Information on reimbursement for PI panel membership |  |  |
| 2 11/09/2020 | Work package 1, review 1, emerging findings about outcome measures, including InterRAI<br>Work package 1, review 2 | Few examples of outcome measures which incorporate representation from families<br>InterRAI as a long list, there should be attention on how the factors interact for individuals. Categories broad and may not pick up on nuance e.g. for a person quite ill with dementia<br>The functional implications should be emphasised – e.g. potential for social isolation if sensory needs not met.<br>Much of this information currently already collected, but time consuming on paper, not all easily shared with families, and not always used to support responsiveness to change in resident condition.<br>For SK to link with software providers<br>DACHA has useful role in helping establish purpose of a MDS. This could help consistent information be collected across care homes | The key points informed the interpretation of findings and the discussion in the paper reporting the literature review of outcome measures used in care home research, in which the following points were made:<br>1) outcome measures that are used in research are not often used in the day to day life of care homes. 2) common research outcome measures, specifically Barthel Index, were viewed as outdated as care homes often routinely collect a wider range of data about residents. 3) residents can have day to day fluctuations in outcomes, and most research tools only measure outcomes at a single time point so may not collect an accurate picture of residents. |
|  |  |  | DACHA team wrote and published 'Developing a minimum data set for older adult care homes in the UK: exploring the concept and defining early core principles' |
| 3 05/02/2021 | Terms of Refence and Agreed ways of working<br>DACHA project and purpose of an MDS, in context of other practice initiatives | Agreed<br>Need for an MDS to capture individual functional needs, not simply scales<br>That PI can contribute voices of residents, family carers and care home staff to development of MDS<br>An MDS must replace other data recording, not add to it – ask care home managers what they collect regularly, what is used.<br>To have value MDS must provide feedback to care home managers<br>Algorithms to flag e.g. deterioration would be valuable<br>Data must be held securely<br>MDS should include personal preferences<br>MDS needs to be easily accessible, used, with staff trained in using it, may need to have requirement for regular data entry.<br>Critical for an MDS to have integration with NHS data<br>Transferrable between care homes if a person moves care home | DACHA team wrote and published 'Developing a minimum data set for older adult care homes in the UK: exploring the concept and defining early core principles' |
| 4 07/05/2021 | WP3 Findings from realist review of uses of minimum data sets | Importance of frequent data entry for best use of MDS in supporting care – staff need understanding in order to have ownership<br>Ideally MDS should facilitate 2 way communication between care home and family<br>MDS should enable efficient responses for care homes to requests for data<br>MDS should give a real sense of the whole person, incorporating wellbeing and mental health as well as physical health<br>MDS should facilitate resident involvement in data collection<br>Barthel scale is physically focussed, insensitive to change, seen as outdated, but is used sometimes in care homes to calculate staffing needs.<br>Of the 400 tools used in research, only MMSE and Barthel recognised by panel members<br>Reflection on the extensive demands on care homes to share information, with much duplication<br>Examples of information manager chooses to collect to help monitor individual wellbeing and care provision<br>Examples of data provided to CQC but no feedback on performance compared to other homes<br>Encourage responses from direct care staff<br>Incorporate questions about data for wellbeing and mental health | Impact funding sought to develop accessible messages to care staff about principles of MDS and their key role |
|  | WP1 literature review 1, to inform panel how their input informed the review, and the results.<br>List of types of data collected currently in care homes compiled by panel members – discussion of utility/what would be useful to collect<br>Work package 3, survey of care home managers about data collected | Incentivise completion – e.g. offer training/information back from university for care staff | Informed thinking about whether outcome measures used in research measure what they aim to measure and whether they measure what is most important to residents, family and friends and staff.<br>Influenced reporting in the paper reporting the literature review, measures used in research are frequently not relevant to everyday life in care homes, and don't take account of wellbeing.<br>Development of an infographic to share this message widely to increase general understanding<br>Survey questions informed by list drawn up by panel members<br>Questions added addressing wellbeing and mental health. Increased emphasis on mood and perspectives of relatives in the survey.<br>Draft of survey shared with panel members for further comment. |
| 5 06/08/2021 | Open item – what panel members think DACHA should be considering in relation to resident's information<br>Infographic to show data sharing issues – to | Recording information about diversity of residents, including e.g. ethnicity and sexual orientation, so that outcomes for different groups can be seen<br>MDS needs to be able to develop over time<br>Sensitive prompts to consider detailed unmet needs.<br>Prompts for identifying change in resident's condition<br>Prompts for contact with family<br>Should represent 2 direction information flow<br>CQC and safeguarding should be included<br>Reduce text in infographic<br>Provide brief information for involved residents of information stored about them, as context for discussion<br>Use pictures and short video as well as text<br>Be aware of communication needs of involved residents<br>Use photo library from Centre for Ageing Better | Amendments made to infographic which was then shared via DACHA website |
|  | communicate widely the insights from previous panel discussions (see meeting 4)<br>Developing the public involvement with care home residents<br>Feedback to Panel about changes made to Work Package 3 staff survey in light of their input<br>Invitation to additional meeting with DACHA team member LI to inform development of a study to find out residents' and relatives' priorities for research using trials archive<br>Invitation to join DACHA study Facebook<br>Invitation to contribute to a project about data sharing, care homes and GP practices | Panel members happy to be emailed about this project | These points were incorporated into the activity pack used to facilitate resident public involvement<br>Panel discussion prompted discussion in the project team about who's perspectives are incorporated in an MDS; resident, relative, provider, other?<br>This separate project team contacted panel members. |
| 6 15/10/2021 | WP4 workshop re: data linkage | Data collection already happening in care homes. Quality of data collected will depend on whether it has value to the right people<br>Need to identify who has access to the data being collected. How do we ensure it's being used to improve resident care. Also keen for residents and family carers to be able to access collected data<br>Care home staff want to know more about hospital admissions and how to balance min and max data sets<br>Data safety is paramount. Panel members happy with WP4's plans re: data safety<br>Questions about how many care homes are using digital systems | WP4 shared learning with rest of DACHA team (esp. WP5)<br>WP4 decided to return to ensure PI were involved when developing their analysis plan.<br>A "next step" documented in the minutes was to explore option of providing a plain English privacy statement on the DACHA website (but unclear if actioned). |
|  | Update about PI with care home residents (first round of activities in with two APs) | Need to explore ways to make sure people living with dementia are included (and to consider how other projects have done this)<br>To consult family members with PoA<br>Positively received and panel members happy care homes were tailoring participation to individuals' wishes. | Could be answered via BH's survey.<br>PI team incorporated feedback from APs and put in options to help involved people LWD (e.g., a range of activities to choose from, prompt cards, flexibility in how activities were run).<br>All activities kept optional |
| 7 05/11/2021 | Feedback from DACHA survey re: findings (what data care homes are collecting, perspective on data sharing and an MDS)<br>DHSC provider data set<br>Other | Questions about the care homes that participated (how representative they were of overall). Keen for more information re: analysis and context (e.g., to get insights on why some responding homes did not collect NHS numbers). Some surprises at what some care homes do not collect and how this may impact on care. Questions about how complete the care homes' data is.<br>Feeling that survey focused more on resident health than wellbeing. Quality of life is missing<br>Widespread use of digital technology surprising,<br>Important that data sharing is two-way. Need for context with data, staff training, supportive use of league tables.<br>MH positive re: contacts made between DACHA and Skills for Care Workforce Intelligence, with member of SFC joining the DACHA steering committee. | Consideration of how to present survey findings in a nuanced way.<br>Quality of life measures to be included in the MDS.<br>Promoting this message e.g., through the infographic and public resources/Plain English summaries |
| 8 04/02/2022 | WP5 overview and discussion of study recruitment<br>Data items in the MDS (which outcome measures best capture wellbeing and quality of life)<br>Update re: Study Within A Project (SWAP) about domiciliary care, offer to be involved in PI for this | Information sheet - difficulty of balancing ethics committee's need for technical language and residents still being able to understand it. Suggested amendments e.g., to add how long data would be held. Importance of having different options e.g., easy read<br>Looked at ASCOT, QUALIDEM and ICECAP-O. Liked ASCOT but ?missing sense of overall wellbeing, liked QUALIDEM but is long. Need to consider a) the types of care homes taking part (and if findings generalisable) and b) that staff may rate QoL more highly than a family member.<br>Residential care homes and family members should be involved in data interpretation.<br>Members of the panel volunteered to be involved | Recruitment materials amended (information sheets made clearer, flowcharts added), easy read options included<br>Incorporation of feedback into consultation re: measuring QoL. Discussion of how SWAP can explore some of issues mentioned.<br>WP5 said would try to return for this. Also to share benchmarked data with care homes in their area.<br>Some panel members joined the Study Within A Project (SWAP) |
| 9 06/05/2022 | DACHA consultations<br>WP4: Learning and actions from last panel | Panel asked to help trial consultation survey<br>Keen for two-way flow of information will relatives but acknowledge goes beyond DACHA's remit. However could be a recommendation (future-proofing)<br>Discussion re: trusted data sources. Keen for inclusion of district nursing records. | Feedback incorporated into survey design<br>This recommendation has been communicated at conferences and in other outputs<br>Feedback about trusted data sources taken into account by WP4. Inclusion of community service utilisation re-ranked as high priority for MDS capture. |
|  | WP4: Current challenges – data sharing agreements<br>Infographic feedback | Access to information from GPs is problematic for care homes. | WP4 ensured community health records linked in Pilot MDS. Community health data collected and analysed in pilot.<br>Panel thanked for their help, still able to feedback via email if desired.<br>Infographic uploaded to website. |
| 10<br>05/08/2022 | WP5: update and discussion (how to engage with residents and families about DACHA, maintaining care home engagement over time)<br>Update of PI activities with residents<br>Facebook re-launch | Discussion of strategies, e.g., open days, posters, linking recruitment to monthly resident reviews, use of newsletters to keep engaged.<br>Panel advised on how to frame exploring QoL.<br>Panel asked to look at Facebook page and feedback. Some mixed feelings about how useful/appropriate it would be. | WP5 team used a newsletter to keep in touch with care homes.<br>Advice taken into account when designing activity pack.<br>Facebook page amended but eventually taken down. |
| 11<br>04/11/2022 | WP4: Analysis protocol (what is already known, what would be useful to know)<br>Feedback from QoL consultation<br>WP5: Recruitment challenges | Interest in use of data for constructive benchmarking, outcomes that would be interesting (e.g., pressure ulcers, UTIs), importance of context with data – e.g. the particular population of any one care home, staff skill mix and level of training. Need to think about who is receiving the data and their understanding of it. Pros and cons of benchmarking.<br>Panel fed back about how they would like to learn about the findings (infographic or slides, inclusion of more detail in an appendix).<br>Discussion of how to maximise recruitment via family members (when residents cannot consent) as relatives are asking what data would be taken, why DACHA needs it, how their relative would benefit from taking part. Panel suggest making sure activity coordinators have information to pass to relatives, assurance of confidentiality, information meeting for relatives led by care home manager, importance of personal contact from a researcher. | WP4 mindful of feedback when completing their analysis.<br>QoL consultation is summarised in a report of the national consultations on the DACHA website<br>Researcher reflected on the feedback which reinforced the range of approaches being used. |
| 12<br>03/02/2023 | Implementation<br>Reflective exercise | Discussed factors that would potentially affect implementation. Importance of speaking to managers and care staff, also considering the MDS with respect to the “bigger picture”.<br>Discussed the panel’s experiences of being part of DACHA and any feedback. | Reinforces decision for WP5/implementation team to be interviewing and gathering feedback from care homes.<br>Added action points to panel minutes. Keen to see a paper and/or report about how the panel has influenced DACHA (AK focusing paper on this, KM gathering impact data). Started having brief bullet point updates about ongoing work packages at start of each panel (to help panel members to keep track over time). |
| 13<br>05/05/2023 | WP5: Update and MDS preview | Discussed potential differences in people in different staff roles completing outcome measures on behalf of residents.<br>Discussed difficulties of participating care homes completing all data entry in one locality involved in the study and how best to address this. Concern raised by panel of impact on care homes of pressure to complete data entry for the DACHA pilot | Question asking about job roles added to interviews about how care staff complete outcome measures (this fed back to PI panel in meeting 15).<br>WP5 team followed suggestions of the panel in resolving difficulties of data entry completion (offered online debriefs with managers, sent email to managers apologising for issues, made a roadmap for managers, newsletter to update care |
|  | WP1: Discussion of findings of review 2<br>Discussed contributing to writing activities | Discussed wastefulness re: number of outcome measures used in research, how it is unclear how or why they are selected for use. The panel expressed an interest in any end of life measures included.<br>Several panel members expressed an interest. | homes, offered contact via email and phone). (Feedback to panel on this in meeting 15)<br>Comments included in Plain English version of the review and shared with lead reviewer. Attempted to publish the Plain English version but was not picked up – put on website.<br>MK and EA wrote paper, published with KM. Panel members invited to contribute to write up of Panel public involvement in the DACHA study. |
| 14<br>29/06/2023 | Panel not completed – illness of panel member. |  |  |
| 15<br>04/08/2023 | WP4: Analysis update<br>WP5: Feedback from PI team about how feedback on care home pressures was actioned (see meeting 13). | The panel asked questions and discussed what they would be interested in learning from the data (such as how medication is used) and the complexities of interpreting data (e.g., frailty scores). Panel interested in being involved in interpretation of the data. Panel asked if data would be collected on DNAR and on end of life plan (highlighting the difference between these)<br>Panel emphasise importance of staff being able to use the information in an MDS to improve care for individuals.<br>The panel were pleased that this had been actioned. | Feedback incorporated into analysis plan. WP4 event (March 2024) re: data analysis and interpretation.<br>WP4 checked if this could be added to GP data request. Information on discussion of preferred place of death was obtained from community services data set in the pilot MDS, and discussed in publication of the analysis of pilot data. Impact funding to develop accessible information for care staff about their critical role in data (entering and use of data). |
| 16<br>03/11/2023 | Suggestions to enhance care home recruitment to SWAP<br>WP2: Update on VICHTA follow-on study<br>Discussion of an additional panel meeting | The panel made several suggestions e.g., posters up in staff rooms with offer of voucher, easy link to make contact and one to one interviews rather than focus group.<br>The panel felt researchers should be able to submit questions. Discussion of potential uses of VICHTA data.<br>The panel expressed interest in a final event in Spring 2024. The panel reflected on how members' involvement in, or understanding of, research has increased since becoming involved in DACHA. | This approach was used by the SWAP and also when recruiting care home staff for the 3<sup>rd</sup> DACHA consultation.<br>VICHTA researcher to offer panel members chance to respond to VICHTA consultation when live, summer 2024.<br>A panel event was arranged for Spring 2024. Panel reviewed and comments on PI section of final report. |
| 17<br>01/03/2024 | Reflection on participation in DACHA PI panel<br>Emerging findings from WP5, pilot of MDS in care homes | Discussion on how the QoL measures performed in the pilot, how the information might be used to inform care, tensions between standardisation versus tailoring of e-records software for care homes.<br>Discussion on how answering QoL questions had changed care staffs' perceptions of what was important to individuals. | Summary of impact of PI on DACHA study sent out to PI panel members. |
|  | Feedback from principle investigator of DACHA on impact of PI |  |  |

### How the involvement influenced the DACHA project

Influences of the involvement were extensive, pervasive and dynamic, as researchers’ appreciation of the care home data context deepened, PI contributors developed understanding of different perspectives and of research approaches, and with the iterative nature of the involvement, influence early on in the project had ongoing effects later. Key themes are listed and then discussed below.

### Themes

1. *Deepened understanding of the data environment in care homes*
2. *Influence on the pilot MDS*
3. *Aiming for best research practices with care homes*
4. *Personal/professional development for public involvement members*
5. *Expectations of the project*

#### 1. Deepened understanding of the data environment in care homes

Data that care homes were expected to provide to other organisations, and the information about their residents that they did, or didn’t, have access to was a key topic in this project, and this theme was discussed regularly in the PI panel in varied contexts. Discussions were iterative, with topics returning to the agenda of subsequent panel meetings as researchers engaged with input from the panel, came back to the panel to report how they had responded to the input, and discussed implications for the next stage of work.

Early in the project, a literature review of measures or instruments used in care home research was conducted (3) with the aim of discovering if any of the measures would be useful as part of a minimum data set. The emerging findings of this review were discussed with the PI panel, revealing that existing measures gave insufficient attention to mental health or wellbeing. Any measures used should be sensitive to change over time, with different aspects of needs coming to the fore at different times in the trajectory of a person’s stay in a care home. Discussion with the panel members drew attention to how little these examples of measures used in research included representation from families, who are an important source of information about residents. Existing measures were thought to be insensitive, lacking the detail and range of information now routinely collected by care homes to inform their care of residents. As the family members and care staff on the panel engaged with the discussion of what might be in an MDS, they illuminated current usual practice of monthly wellbeing reviews, although not necessarily shared with families. The panel discussion was reflected in the report of the literature review, in particular that there is little relationship between outcome measures used in research and routine data recording in care homes, research measures appear outdated in relation to information recorded in care homes and insensitive to day to day fluctuations for residents (3).

As researchers and panel members listened to each other and worked to reach shared understandings about the purpose of a minimum data set in care homes researchers found it difficult to explain the parameters for data with potential for inclusion in an MDS. There were differing interpretations both among the research team and with and between the PI Panel members. The thoughtful questioning and challenging discussions prompted the DACHA team to negotiate, agree and propose a definition of and purpose for an MDS (29).

PI Panel members emphasised the desirability of integration between care home records and NHS data, transferability for a person moving from one care home to another and usability for staff. Detailed accounts of the day-to-day realities of dealing with data in care homes were crucial to the research team appreciating the complexities of the demands placed on care home staff to provide data to other agencies. Very similar data is required by many different stakeholders, and different departments in the same stakeholders, in different formats, leading to duplication of effort. The experience for care homes is of providing data but getting nothing back in the way of analysis or feedback on how their outcomes related to those of other care homes of similar size and locality. Three members of the panel (2 managers and 1 senior carer) completed a list of all the types of data that they recorded regularly and this was used in the design of a survey sent out to care home staff (5). With input from the panel an infographic was designed to communicate the data demands on care homes (30).

It was clear that an MDS should draw on existing data and not add to the burden of care homes. To add value, and therefore to motivate implementation, an MDS must provide feedback to care home managers on the performance of their care home, to feedback to teams and drive improvement. The PI panel were aware of the importance of the work and increasing national focus on care homes and data as a consequence of the impact of the COVID-19 pandemic and policy responses.

#### 2. Influence on the pilot MDS

### Quality of life

The family members, staff and managers in the PI Panel were clear that an MDS should give a real sense of the whole person, incorporating wellbeing and mental health. The discussions between the DACHA researchers and the PI panel were iterative: having fed into the development of an online survey for care home staff (see section above), the results from the survey were fed back to the panel and discussed in order to inform the analysis. This drew attention to the dearth of information being recorded in practice about quality of life. Further analysis revealed that information about quality of life was recorded by fewer than a third of respondents to the survey (5). The impetus from the PI Panel was to push the developing DACHA MDS beyond international examples to incorporate quality of life. Further work together between the DACHA researchers and PI Panel members included discussion of the pros and cons of a selection of measures that could potentially capture quality of life as part of an MDS. The PI panel contributed to the development of activities that could be used to facilitate public involvement from care home residents about quality of life and how it could be captured (27), and their views also informed the development of a consultation with wider stakeholders (7).

In response, the DACHA team reviewed quality of life measures that could potentially be used. There was consultation with stakeholders on utility and usability of a number of measures and a short list of possible quality of life measures were discussed with the PI panel. As a result these measures were included in the pilot MDS that was trialled in 45 care homes with 996 residents (ref) in Work Package 5 of DACHA (WP5). Three of the four measures piloted were found to have acceptable psychometric properties (31) and used to better understand the factors associated with different constructs of residents’ QoL (for example, emergency hospital admissions) (32).

### Community health data

The Health Foundation (THF) led work package 4 in the DACHA project, bringing expertise in data linkage and analysis of sets of routine health data. While simple in conception, the execution is complex both technically and in relation to governance processes. THF DACHA team members drafted accessible presentations, discussed and refined these in collaboration with DACHA PI research team members and met the PI Panel four times (Panel meetings 6, 9, 11 15). PI Panel members were able to develop an understanding of a complex and technical research approach so that they could contribute their views and get feedback on how their input had influenced the research. The PI Panel were excited by the potential of linking individual care records with health records, and particularly emphasised the value of linking information on community health care including district nursing and community rehabilitation services. Panel members working in care homes commented on the strength of district nursing records as a source of reliable information about residents’ health and input from health services. This resonated with public involvement with care home residents (27) who put priority on better information about their appointments with health professionals. The DACHA RMT met face to face to agree principles and priority information for a minimum data set (May 2022) and the PI team members advocated for community health information to be included in the pilot MDS through data linkage, as this was prioritised by PI Panel members and residents. In response the pilot MDS included mean number of community services appointments overall, and for each of five priority services: speech and language therapy, continence, district nursing, podiatry, and community rehabilitation. All were reported over one year and could be summarised across different subgroups, for example by resident or care home characteristics, to understand variation. Information collected during such appointments was not accessed.

#### 3. Aiming for best research practices with care homes

Involvement of a family carer and a director of the NCF from the early stages of project development and writing the funding bid helped to ensure that the practical implications of carrying out the research for care homes and for the people living in them were kept central to discussions. The DACHA project aimed to create new ways of working and doing research in and with care homes, so that the outputs benefit not only researchers but also residents. The research team’s awareness of the demands on care homes and the workload for staff and managers was sharpened by the panel discussions over the project. Participating in research would bring demands over and above day to day practice, which was still recovering from the impact of COVID-19, and the hours of participation should be made clear to care homes, as transparency aids the homes’ planning and commitment. There was a sense that the needs of the care home should be central and the demands of taking part in the research should work around this. A critical point in the timings of the research project challenged this value leading to difficult discussions both in the PI Panel and the DACHA team. The pilot of the MDS involved care homes completing additional measures at two time points for each participating resident in their home, in addition to routinely collected data for these residents being extracted. There was a deadline for completing the measures in order for the e-record software providers to extract the data. There was a miscommunication, some care home managers weren’t informed that there was a request for additional measures to be completed and the deadline for these.

The researcher dealing with the consequences of the missed information was negotiating a course balancing the evident stress of care home managers and staff when approached to complete data entry in a short space of time and the demands for the viability of key aspects of the research study. When the researcher presented this as part of an update on progress of the study to the PI Panel, panel members expressed disappointment that, despite their involvement and contributions throughout the study and the expressed wish of the study to work well with care homes, past poor practices experienced by some panel members in other research projects had been repeated. The respectful but challenging exchange in the PI Panel meeting, which a panel member thought benefitted from experienced chairing to ensure all views were heard, respected and understood, did develop an action plan to mitigate the impact on the care homes involved. This included offering online debriefing sessions to care home managers, a communication from the study lead apologising for the issues, and the offer of contact by email or phone for care home managers with researchers. A ‘road map’ of future dates was suggested by a care home manager at a participants’ debriefing session and sent to all participating care homes.

Most of the affected care homes strove to complete the additional measures in the short time frame believing that the project will benefit residents in the long-term. Others, already stressed by issues other than the research, withdrew from the study (then or just after). A panel member reflected on the importance of endings and the impact on any future research participation for these homes. The local research nurse contacted the withdrawn care homes to understand learning and keep communication channels open for future research.

The uncomfortable position for the researcher in the discussion at the PI Panel as the spokesperson for the conduct of the fieldwork, was discussed at a face-to-face RMT meeting. For some team members the practical problems faced by the project were seen to make unavoidable any additional pressure put on participating care homes. Indeed this was also respecting the individual residents in those participating homes who had given consent for their records to be used in the study. Others argued that it was important for the project to act consistently with the value of giving care homes a voice in the research both through the PI Panel and relations with participating care homes.

Even though we aimed to ensure that the needs of the care home should be central and we had strong PI, undue pressure on care homes can quickly arise (e.g. through a miscommunication).

#### 4. Personal/professional development for public involvement members

The team did not aim for involvement in the panel to be a developmental experience for the members but it was clear from a reflective session held in meeting 12 that it was for some. Panel members described their motivation for getting involved as wanting to make a contribution. Family carers felt they could draw on their experience of having their spouse or parent living in a care home. They were also drawing on other life experiences such as their own current or previous work or volunteering in health or care related settings or local authorities. Some family carer panel members described their contribution as answering questions and saying how things had been for them, saying that researchers and people working in care homes should influence the research more.

One family carer expressed that they had learned from the project, and been helped with using technology (for the online meeting process). They subsequently raised awareness of public involvement while volunteering with dementia groups.

By this 12^th^ meeting, panel members were able to describe initial concerns that taking part in the panel would raise uncomfortable differences in perspectives. One family carer described being fearful that they would be ‘too negative’ as a panel member because they had not been able to find good care for their relative. This person worked in an organisation aiming to support and promote social care so was wary of the potential conflict with that role if they spoke negatively about care, but reflected that taking part had sparked ideas for improvement of her organisation’s work. A senior carer described their initial caution in contributing because of the ‘disconnect and lack of understanding’ in society of the work of social care. This person was concerned that the research plan would be unrealistic. However, they described the ‘morale boost’ of the unexpected opportunity to connect with a group of people who, from different perspectives, cared deeply about the subject. They had considered things that wouldn’t have occurred to them, which had informed and improved their work as a carer. Care home managers in the panel valued hearing thoughts and reflections of family members who could be frank and open in this different context, and this influenced their practice. Two members of the panel published an article aimed at care home staff about their experience on the panel, and the career opportunities this led to which included winning research funding as a co-applicant and working for NIHR (28) and a career path blending practice, research and implementation.

#### 5. Differing Expectations of the project

Some issues of great importance to PI Panel members were not necessarily within the scope of the project. An early example was whether the pilot MDS being developed in the project would share real time data with families. Panel members were cognizant of the huge potential of linkage of individual care and health records for effective care. The scope of the pilot MDS was to create a pseudonymised proof-of-concept linked dataset that could provide useful insights about residents but would not identify individual residents or be directly accessible to CH staff, residents or carers (so therefore couldn’t share any data with families). From the perspective of family members this was too narrow an objective. They stressed the potential to use software to be able to easily find out how their family member was, what they have been doing during the day, without calling care staff away from their work with residents to give information over the phone. Digital care planning systems were evolving and their use became more widespread in care homes during the course of the project. Some of the functionality that panel members thought important in an MDS was becoming available in e-records systems, including family members being able to access information about their relative, and systems flagging certain signs of deterioration in individuals.

There was some frustration from PI contributors about the pace of setting up public involvement activities, the timing and format of reporting meetings back to panel members and the limitations of an on-line rather than a face-to-face meeting. The Principal Investigator (CG) and research team responded with telephone conversations and in-person meetings with concerned individuals and agreed action plans to try to resolve issues. The original family member co-applicant chose to leave the project, giving the reason that their time could be better used elsewhere.

The PI team aspired for more PI Panel involvement in data analysis and interpretation than was achieved as the complexities of study recruitment, data governance between organisations for data linkage and data extraction limited the scope of the MDS and the amount of time available for analysis. Panel members were interested to contribute to writing up the work of the panel and have contributed to this paper.

## Discussion

The DACHA project set out to keep the concerns and priorities of people living and working in care homes informing all stages of the project and we argue that the examples of the content of the pilot MDS show that this has been achieved. We aimed to use a democratic approach, valuing different knowledges. There is evidence in reflections from the panel that members did feel that their knowledge and experience was valued and heard.

However such an approach is fragile and can be challenged by the contractual obligations of completing a funded research project in the context of unexpected events (33). Fragile democratic relations between the research team and the PI panel members could be argued to have been bolstered in this example by the consistent process of quarterly meetings with dates set when the panel was formed, and by chairing that aimed to developed shared understanding and trust between people attending each panel meeting.

It is of course oversimplistic to characterise relations as simply between PI panel members and the research team as many had more than one role or identity. The research team included members chosen for their link to practice (NCF) and family carer experience. Researchers working for academic institutions also have relevant family or social care practice experience. The research team began with 14 collaborators, then grew substantially as researchers were brought in to work on various aspects of the study, and as additional funding was won to develop particular aspects of the study. Some of the team know each other well and had collaborated in previous research while others were working together for the first time. PI Panel members, both family members and practitioners, brought experience in other roles including research, social care practice and advocacy and volunteering (34). Some developed research knowledge that helped them into other research related roles, meeting the policy agenda for developing social care research capacity (35). These overlapping roles brought shared experience and empathy to the panel interactions, but also as ‘boundary spanners’ the potential for advocating for social care research grounded in practice.

These multiple roles and identities also underpin complex power relations. Russell et al (20) argue that how such power relations play out and the interests served by empowerment should be considered in the public involvement research agenda. Reflections in the RMT meeting and in the PI Panel have touched on these issues. Notably the concern and sense of responsibility expressed by PI Panel members either initially sceptical that a research project could be ‘realistic’, or struggling to reconcile a felt professional responsibility to advocate for the care sector with personal experience of poor practice.

We argue there have been ‘soft’ effects from the PI process that are hard to capture but are important impacts on growing a social care research practice. Researchers’ understanding of and attitudes towards residents and care home staff have benefitted from a deeper understanding and appreciation their situation. Many of us have completed the project having learned far more than can be wrapped up and capitalised on in this project. Such is the nature of project-based research in an academic and funding context that was not set up with the care sector in mind and could usefully adapt quickly so that the care sector gets the relevant developing evidence base and potential to influence policy.

## Conclusion

The DACHA project shows what can be achieved with integrating public involvement throughout a project to powerful effect, with leadership and commitment demonstrated from the Chief Investigator and with sufficient resources designed into the project. The public involvement not only informed the MDS but also deepened our understanding of the context in which we were working and provided both accountability and support when there were issues. The public involvement exemplified a ‘social practice of dialogue and learning between researchers and the public’(20). In such practice it is likely there will always be challenges in developing shared understandings and expectations. Next steps of development and implementation of an MDS for care homes should build on such relations, incorporating relevant knowledge and experience, in order to minimise negative unforeseen consequences. There is a need to tap the deep knowledge in practice by spanning boundaries between research and practice, and rapidly enhance practitioner/research in social care.

## Conflict of interest statement

No conflicts of interest reported by authors. Emily Allison now works for the NIHR (but did not at outset of the project). This was not considered to be a conflict of interest by the DACHA team or the NIHR because her work is in an entirely separate department to the funding stream for this project. Since commencing employment at the NIHR she has not accepted remuneration from DACHA.

## Data availability statement

Data sharing not applicable to this article as no datasets were generated or analysed during the current study

## Ethics approval statement

The DACHA project received ethical approval for distinct elements of the research project.

- Work Package 2: received ethical approval from Health, Science, Engineering & Technology ECDA – University of Hertfordshire (HSK/SF/UH/04185)
- WP3 national care home survey: received ethical approval from Health, Science, Engineering & Technology ECDA – University of Hertfordshire (HSK/SF/UH/04301)
- WP5 care home pilot: received ethical approval from the London Queen’s Square Research Ethics Committee (22/LO/0250).
- National consultation 2022: received ethical approval from Health, Science, Engineering & Technology ECDA – University of Hertfordshire (HSK/SF/UH/05009)
- National consultation 2023-24: received ethical approval from Health, Science, Engineering & Technology ECDA – University of Hertfordshire (HSK/SF/UH/05487)

Ethical review was not sought for this analysis of the PI process as it is not required for public involvement activity.

## Funding statement

This study/project is funded by the National Institute for Health and Care Research (NIHR) Health Service Research and Delivery programme (HS&DR NIHR127234) and supported by the NIHR Applied Research Collaboration (ARC) East of England.

The views expressed are those of the author(s) and not necessarily those of the NIHR or the Department of Health and Social Care.

## Acknowledgements

We gratefully acknowledge the public involvement contributors who made an invaluable contribution to the DACHA project, and the DACHA team members who all committed to working thoughtfully with the public contributors. Thanks to Sue Stirling and Stacey Rand for helpful suggestions on the paper, and to Priti Biswas for early work with the DACHA PI team.

---

GRIPP 2 Long Form for reporting public involvement(36)

GRIPP 2 Long Form for reporting public involvement(36)
| Section and topic | Item | Reported on page No |
| --- | --- | --- |
| Section 1: Abstract of paper |  |  |
| 1a: Aim | Report the aim of the study | 1 |
| 1b: Methods | Describe the methods used by which patients and the public were involved | 1 |
| 1c: Results | Report the impacts and outcomes of PPI in the study | 1 |
| 1d:Conclusions | Summarise the main conclusions of the study | 1 |
| 1e: Keywords | Include PPI, “patient and public involvement,” or alternative terms as keywords | 1 |
| Section 2: Background to paper |  |  |
| 2a: Definition | Report the definition of PPI used in the study and how it links to comparable studies | 2-3 |
| 2b: Theoretical underpinnings | Report the theoretical rationale and any theoretical influences relating to PPI in the study | 2-3 |
| 2c: Concepts and theory development | Report any conceptual models or influences used in the study | 2-3 |
| Section 3: Aims of paper |  |  |
| 3: Aim | Report the aim of the study | 2 |
| Section 4: Methods of paper |  |  |
| 4a: Design | Provide a clear description of methods by which patients and the public were involved | 4-6 |
| 4b: People involved | Provide a description of patients, carers, and the public involved with the PPI activity in the study | 4-6 |
| 4c: Stages of involvement | Report on how PPI is used at different stages of the study | 4-6 |
| 4d: Level or nature of involvement | Report the level or nature of PPI used at various stages of the study | 4-6 |
| Section 5: Capture or measurement of PPI impact |  | 6-12 |
| 5a: Qualitative evidence of impact | If applicable, report the methods used to qualitatively explore the impact of PPI in the study | 6-12 |
| 5b: Quantitative evidence of impact | If applicable, report the methods used to quantitatively measure or assess the impact of PPI | NA |
| 5c: Robustness of measure | If applicable, report the rigour of the method used to capture or measure the impact of PPI | NA |
| Section 6: Economic assessment |  |  |
| 6: Economic assessment | If applicable, report the method used for an economic assessment of PPI | NA |
| Section 7: Study results |  |  |
| 7a: Outcomes of PPI | Report the results of PPI in the study, including both positive and negative outcomes | 6-12 |
| 7b: Impacts of PPI | Report the positive and negative impacts that PPI has had on the research, the individuals involved (including patients and researchers), and wider impacts | 6-12 |
| 7c: Context of PPI | Report the influence of any contextual factors that enabled or hindered the process or impact of PPI | 6-12 |
| 7d: Process of PPI | Report the influence of any process factors, that enabled or hindered the impact of PPI | 6-12 |
| 7ei: Theory development | Report any conceptual or theoretical development in PPI that have emerged | 13 |
| 7eii: Theory development | Report evaluation of theoretical models, if any | 13 |
| 7f: Measurement | If applicable, report all aspects of instrument development and testing (eg, validity, reliability, feasibility, acceptability, responsiveness, interpretability, appropriateness, precision) | NA |
| 7g: Economic assessment | Report any information on the costs or benefit of PPI | NA |
| Section 8: Discussion and conclusions |  |  |
| 8a: Outcomes | Comment on how PPI influenced the study overall. Describe positive and negative effects | 6-12 |
| 8b: Impacts | Comment on the different impacts of PPI identified in this study and how they contribute to new knowledge | 6-12 |
| 8c: Definition | Comment on the definition of PPI used (reported in the Background section) and whether or not you would suggest any changes | 2-3 |
| 8d: Theoretical underpinnings | Comment on any way your study adds to the theoretical development of PPI | 13 |
| 8e: Context | Comment on how context factors influenced PPI in the study | 6-12 |
| 8f: Process | Comment on how process factors influenced PPI in the study | 6-12 |
| 8g: Measurement and capture of PPI impact | If applicable, comment on how well PPI impact was evaluated or measured in the study | NA |
| 8h: Economic assessment | If applicable, discuss any aspects of the economic cost or benefit of PPI, particularly any suggestions for future economic modelling. | NA |
| 8i: Reflections/critical perspective | Comment critically on the study, reflecting on the things that went well and those that did not, so that others can learn from this study | 6-13 |

PPI=patient and public involvement

## Notes

### Competing Interest Statement

The authors have declared no competing interest.

## References

1. Towers A-M, Gordon A, Wolters AT, Allan S, Rand S, Webster LA, et al. Piloting of a minimum data set for older people living in care homes in England: protocol for a longitudinal, mixed-methods study. BMJ Open. 2023;13(2):e071686.

2. Musa MK, Akdur G, Brand S, Killett A, Spilsbury K, Peryer G, et al. The uptake and use of a minimum data set (MDS) for older people living and dying in care homes: a realist review. BMC Geriatrics. 2022;22(1):33.

3. Kelly S, Cowan A, Akdur G, Irvine L, Peryer G, Welsh S, et al. Outcome measures from international older adult care home intervention research: a scoping review. Age Ageing. 2023;52(5).

4. Peryer G, Kelly S, Blake J, Burton JK, Irvine L, Cowan A, et al. Contextual factors influencing complex intervention research processes in care homes: a systematic review and framework synthesis. Age and Ageing. 2022;51(3).

5. Hanratty B, Wolters AT, Towers AM, Spilsbury K, Meyer J, Killett A, et al. Data Collection in Care Homes for Older Adults: A National Survey in England. Journal of Long-Term Care. 2023:288–96.

6. DACHA. Feedback on DACHA study’s 2021 consultation events http://dachastudy.com/wp-content/uploads/2021/10/Report-DACHA-consultation-2021.pdf: DACHA; 2021 [

7. DACHA. Quality of Life Consultation Feedback Report [Internet] http://dachastudy.com/wp-content/uploads/2022/12/DACHA-2022-Consultation-report-FINAL-.pdf2022 [cited 2023 19 Jan 2023].

8. DACHA. DACHA Final Consultation on Minimum Data Set - Feedback Report [Internet] http://dachastudy.com/wp-content/uploads/2024/05/DACHA-consultation-feedback-report-2024-v3.pdf2024 [

9. Gordon AL, Franklin M, Bradshaw L, Logan P, Elliott R, Gladman JRF. Health status of UK care home residents: a cohort study. Age and Ageing. 2013;43(1):97–103.

10. DHSC. A plan for digital health and social care. In: Care DoHaS, editor. https://www.gov.uk/government/publications/a-plan-for-digital-health-and-social-care/a-plan-for-digital-health-and-social-care: Gov.UK; 2022.

11. Shachak A, Buchanan F, Kuziemsky C. When rules turn into tools: An activity theory-based perspective on implementation processes and unintended consequences. Healthc Manage Forum. 2024;37(3):177–82.

12. Ostaszkiewicz J, O’Connell B, Dunning T. Fear and overprotection in Australian residential aged-care facilities: The inadvertent impact of regulation on quality continence care. Australas J Ageing. 2016;35(2):119–26.

13. Burgher T, Shepherd V, Nollett C. Effective approaches to public involvement in care home research: a systematic review and narrative synthesis. Research Involvement and Engagement. 2023;9(1):38.

14. Logan PA, Horne JC, Gladman JRF, Gordon AL, Sach T, Clark A, et al. Multifactorial falls prevention programme compared with usual care in UK care homes for older people: multicentre cluster randomised controlled trial with economic evaluation. BMJ. 2021;375:e066991.

15. Froggatt K, Goodman C, Morbey H, Davies SL, Masey H, Dickinson A, et al. Public involvement in research within care homes: benefits and challenges in the APPROACH study. Health Expect. 2016;19(6):1336–45.

16. Stirrup O, Tut G, Krutikov M, Bone D, Lancaster T, Azmi B, et al. Anti-nucleocapsid antibody levels following initial and repeat SARS-CoV-2 infections in a cohort of long-term care facility residents in England (VIVALDI) [version 1; peer review: 2 approved]. Wellcome Open Research. 2024;9(45).

17. Stocker R, Brittain K, Spilsbury K, Hanratty B. Patient and public involvement in care home research: Reflections on the how and why of involving patient and public involvement partners in qualitative data analysis and interpretation. Health expectations: an international journal of public participation in health care and health policy. 2021;24(4):1349–56.

18. Edelman N, Barron D. Evaluation of public involvement in research: time for a major re-think? J Health Serv Res Policy. 2016;21(3):209–11.

19. Burns D, Hyde P, Killett A, Poland F, Gray R. Participatory organisational research: Examining voice in the co-production of knowledge. British Journal of Management. 2012;25:133–44.

20. Russell J, Fudge N, Greenhalgh T. The impact of public involvement in health research: what are we measuring? Why are we measuring it? Should we stop measuring it? Research Involvement and Engagement. 2020;6(1):63.

21. Frith L. Democratic Justifications for Patient Public Involvement and Engagement in Health Research: An Exploration of the Theoretical Debates and Practical Challenges. The Journal of Medicine and Philosophy: A Forum for Bioethics and Philosophy of Medicine. 2023;48(4):400–12.

22. Strasser BJ, Baudry J, Mahr D, Sanchez G, Tancoigne E. “Citizen Science”? Rethinking Science and Public Participation. Science & Technology Studies. 2019;32(2):52–76.

23. Beresford P, Slasberg C. The Future of Social Care Edward Elgar Publishing; 2023.

24. Banerjee A, Armstrong P, Daly T, Armstrong H, Braedley S. “Careworkers don’t have a voice:” Epistemological violence in residential care for older people. Journal of Aging Studies. 2015;33:28–36.

25. Fricker M. Epistemic justice as a condition of political freedom? Synthese. 2013;190(7):1317–32.

26. Staniszewska S, Brett J, Simera I, Seers K, Mockford C, Goodlad S, et al. GRIPP2 reporting checklists: tools to improve reporting of patient and public involvement in research. Research Involvement and Engagement. 2017;3(1):13.

27. Micklewright K, Killett A, Akdur G, Biswas P, Blades P, Irvine L, et al. Activity provider-facilitated patient and public involvement with care home residents. Res Involv Engagem. 2024;10(1):7.

28. Kelly M, Allison E, Micklewright K. Health and social care research from the frontline: perspectives from care home staff. Nursing and Residential Care. 2023;25(11):1–3.

29. Burton JK, Wolters AT, Towers AM, Jones L, Meyer J, Gordon AL, et al. Developing a minimum data set for older adult care homes in the UK: exploring the concept and defining early core principles. Lancet Healthy Longev. 2022;3(3):e186–e93.

30. DACHA. Where is information recorded when a person in an English care home falls? https://dachastudy.com/wp-content/uploads/2022/08/FINAL-DACHA-pdf.pdf2022.

31. Towers A-M, Rand S, Allan S, Webster L, Palmer S, Carroll RE, et al. Cross-sectional study assessing the feasibility of measuring residents’ Quality of Life in English care homes and assessing the construct validity and internal consistency of measures completed by staff-proxy. medRxiv. 2024:2024.05.20.24307612.

32. Allan S, Rand S, Towers A-M, De Corte K, Tracey F, Crellin E, et al. Factors associated with care home resident quality of life: Demonstrating the value of a pilot Minimum Data Set using cross-sectional analysis from the DACHA study. medRxiv. 2024:2024.05.30.24308190.

33. Richards DP, Poirier S, Mohabir V, Proulx L, Robins S, Smith J. Reflections on patient engagement by patient partners: how it can go wrong. Research Involvement and Engagement. 2023;9(1):41.

34. Forbat L, Macgregor A, Brown T, McCormack B, Spilsbury K, Rutherford A, et al. Negotiating pace, focus and identities: Patient/public involvement/engagement in a palliative care study. Sociology of Health & Illness.n/a(n/a).

35. Pulman A, Fenge L-A. Building capacity for social care research – ways of improving research skills for social workers. Social Work Education. 2024;43(1):60–78.

36. Staniszewska S, Brett J, Simera I, Seers K, Mockford C, Goodlad S, et al. GRIPP2 reporting checklists: tools to improve reporting of patient and public involvement in research. Bmj. 2017;358:j3453.

